# Host Factors Modulate Nirmatrelvir–Ritonavir Efficacy in COVID-19 Patients: A Viral Dynamics Modeling Study

**DOI:** 10.64898/2026.04.03.26350141

**Authors:** Liao Yiyu, Yan Wang, Yuqian Wang, Jingwen Ai, Law Boon Kiat, Derrick Lim, Jiaxin Zhou, Hongyu Wang, Yanpeng Wu, Po Ying Chia, Hoong Kai Chua, Conrad En Zuo Chan, Joshua T. Schiffer, Katherine Owens, Shadisadat Esmaeili, Benjamin J. Cowling, Matthew E. Cove, Hiroki Saito, Liang En Wee, Barnaby E. Young, Tat Ming Ng, Eric Chun Yong Chan, Marco Ajelli, Wenhong Zhang, Hongjie Yu, Keisuke Ejima

## Abstract

Antiviral therapies such as nirmatrelvir–ritonavir are widely used for COVID-19, yet their real-world effectiveness and sources of heterogeneity in treatment response remain incompletely understood. Here, we integrate longitudinal viral load data from a large cohort of SARS-CoV-2 BA.2–infected patients in Shanghai (n=48,243) with a mechanistic within-host viral dynamics model coupled to pharmacokinetic/pharmacodynamic principles to quantify in vivo antiviral efficacy. We estimate that nirmatrelvir–ritonavir reduces viral production by approximately 55% on average. Treatment response exhibits substantial heterogeneity, with higher efficacy observed in vaccinated individuals and reduced efficacy in older adults. Sensitivity analyses demonstrate that the vaccination effect is robust across model specifications, whereas age-related differences depend on assumptions about early viral kinetics, highlighting structural identifiability challenges when analyzing sparse real-world data. These findings provide a mechanistic interpretation of heterogeneous treatment effects and establish a generalizable framework for integrating real-world clinical data with within-host models to inform antiviral optimization and personalized treatment strategies.

## Introduction

Severe acute respiratory syndrome coronavirus 2 (SARS-CoV-2) is the infectious agent responsible for the coronavirus disease 2019 (COVID-19) pandemic, a global health crisis estimated to have resulted in over 14.8 million excess deaths worldwide in 2020 and 2021^1^. SARS-CoV-2 remains a significant public health concern due to the continuous emergence of evolving variants and its associated post-viral syndrome with long-term complications^2^. Therefore, understanding viral load dynamics and its determinants is key since viral load has been shown to affect both SARS-CoV-2 transmissibility and disease severity^3,4^. Consequently, the viral clearance rate has been used as an endpoint in several clinical trials of pharmacological therapeutics for measuring treatment efficacy^5–8^. For SARS-CoV-2, this is generally determined by quantifying the viral load longitudinally through reverse transcription-quantitative polymerase chain reaction (RT-qPCR) in patients with confirmed infections, usually via a sample from the upper respiratory tract (nasopharyngeal or oropharyngeal swab)^9^.

Several antiviral and non-antiviral drugs have been developed or repurposed for treating SARS-CoV-2 infections. One of the most widely used and internationally approved antivirals is nirmatrelvir–ritonavir (Paxlovid). Nirmatrelvir inhibits the main protease (M^pro^), an enzyme essential for viral RNA replication^10,11^. On the other hand, ritonavir is seldom used solely as an antiviral, but functions as a pharmacokinetic booster by slowing the metabolism of nirmatrelvir, thereby maintaining effective plasma concentrations. Although clinical trials evaluating nirmatrelvir–ritonavir, or other antivirals including molnupiravir and remdesivir, have demonstrated treatment efficacy, substantial individual heterogeneity exists, particularly regarding viral load outcomes^10,12,13^.

One important factor that may influence the efficacy of antiviral therapies is the patient’s age. A meta-analysis of viral load dynamics found that viral RNA shedding is prolonged in older adults across a majority of studies, suggesting a negative association between viral clearance and age^14^ — a finding that was supported by results from mathematical modeling^15^. However, how age influences the effectiveness of antiviral treatment remains unclear. In one study, the treatment effect of remdesivir — measured as the ratio of recovery (defined as either discharge from the hospital or hospitalization) rates between treated and untreated patients — was greater in COVID-19 patients under 40 years old than in older patients, suggesting that the antiviral efficacy of remdesivir may decline with age^16^. Similar trends have been observed in other respiratory viral infections; for example, in an influenza trial, older age among oseltamivir recipients was associated with prolonged viral shedding^17^. In addition to antiviral treatment, age may also influence the effectiveness of vaccination. A number of studies on SARS_CoV_2 vaccines have shown that antibody titers tend to be lower and decline more rapidly in older populations, suggesting a weaker and shorter_lived protective response^18–22^.

Motivated by these findings, we aim to examine whether the antiviral effect of COVID-19 treatments on viral load is influenced by patients’ age and vaccination status. To test the hypothesis that nirmatrelvir–ritonavir treatment efficacy varies across patient subgroups, we applied a mathematical model to real-world data (RWD) from SARS-CoV-2 BA.2-infected patients in Shanghai, China. Understanding the main determinants of the variability in treatment efficacy would provide deeper insights into the antiviral mechanisms of these drugs, informing the development of more effective antiviral therapies against SARS-CoV-2 and other RNA viruses, and facilitating the refinement of clinical guidelines for COVID-19 treatment.

## Methods

### Real-world data from hospitalized COVID-19 patients

We leveraged on a large dataset that was collected during the Omicron BA.2 outbreak in Shanghai, China, from March 23 to May 15, 2022, which includes all patients admitted to three designated hospitals or makeshift hospitals^23^. Although viral genomic sequencing was not performed for every individual patient, surveillance data from the early and middle stages of the outbreak confirmed that the circulating SARS-CoV-2 strains in Shanghai during this period were exclusively of the Omicron BA.2 lineage^24^. The dataset comprises 48,243 patients with detailed demographic information, key timelines (e.g., timing of detection, hospitalization, symptom onset, antiviral treatment, clinical progress and discharge), and daily viral load measurements from nasopharyngeal swabs collected from the time of hospitalization or isolation until discharge based on clinical requirements during their hospital stays. It is important to note that during this period, strict contact tracing and mass screening campaigns were implemented in Shanghai. Consequently, a substantial proportion of patients were identified and isolated in the presymptomatic phase or immediately upon viral shedding, distinct from cohorts relying solely on passive symptomatic surveillance.

Viral load was quantified using standard real-time RT-PCR assays targeting the SARS-CoV-2 nucleocapsid (N) and open reading frame 1ab (ORF1ab) genes. To ensure data consistency across the three designated hospitals, sample collection and laboratory testing strictly adhered to the ‘Technical Guidelines for the Collection and Testing of SARS-CoV-2 Samples’ (Protocol on Prevention and Control of SARS-CoV-2, Version 8), issued by the National Health Commission of the People’s Republic of China^25^.

Cycle threshold (Ct) values were converted to viral load (RNA copies/mL) using a linear relationship derived from the standard curve data reported by Zou et al^26^. Based on their reported calibration points, we applied the following conversion equation:

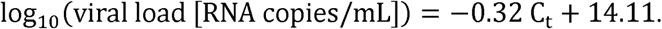

Nirmatrelvir–ritonavir has been frequently prescribed in China as an antiviral treatment for patients with mild to moderate COVID-19, but at high risk of developing severe COVID-19 or its complications^27^. According to the clinical treatment guidelines^27^, we included patients who met the following criteria: (1) symptomatic infection, (2) at least one risk factor for progression to severe disease (e.g., age > 65 years; cardiovascular disease, diabetes, or other chronic illness; body mass index [BMI; kg/m^2^] ≥ 30; or receipt of less than two vaccine doses), (3) age ≥ 18 years, (4) without severe renal impairment (e.g., undergoing dialysis or having received a kidney transplant) or liver disease, (5) received antiviral treatment within 7 days of symptom onset, (6) available data on timing of detection and clinical progression, and (7) did not require oxygen supplementation on the reference date, defined as the date of treatment initiation for the treatment group (treated with nirmatrelvir–ritonavir) and 4 days after detection for the control group (no antiviral treatment), corresponding to the median treatment initiation time in the treatment group. Patients in both the treatment and control groups received standard supportive care in accordance with treatment guidelines. This included the use of antipyretics (e.g., for fever management) and glucocorticoids as clinically indicated. However, the control group was strictly defined as individuals who did not receive nirmatrelvir–ritonavir or any other targeted antiviral therapies (e.g., molnupiravir, remdesivir), which may influence the viral dynamics, during the study period. Patients with fewer than three detectable viral load measurements were excluded from the analysis.

### Retrospective cohort construction

In most observational studies, the treatment is not randomly assigned, which can introduce bias^28^. To mitigate this, we performed propensity score matching^29^. This technique balances potential confounding factors between the treatment (nirmatrelvir–ritonavir) and control (no antivirals) groups by estimating each patient’s probability of having received treatment based on their observed covariates. We estimated propensity scores using the classification and regression tree (CART) algorithm^30^, a decision tree model, which naturally accommodates non-linear relationships and higher-order interactions among covariates. The propensity model was conditional on the age group (18–64 years or ≥ 65 years), sex (male or female), number of comorbidities (0 or ≥1), vaccination status (less than 2 doses, 2 doses, and 3 doses), and initial viral load (defined as the log__ RNA copies/mL measured at the time of detection). Comorbidities included diabetes, hypertension, cardiovascular disease, hematologic disorders, kidney disease, neurological conditions, pneumonia, malignancy, and rheumatism. Although three types of vaccines were administered (i.e., inactivated, adenovirus-vectored, and protein subunit), they were treated as a single category in our analysis. To ensure that the treatment and control groups were comparable, we used optimal pair matching^31^. Unlike greedy matching algorithms, optimal matching minimizes the sum of the absolute distances of propensity scores across all matched pairs in the entire sample, thereby producing the best overall balance without the substantial data loss associated with direct matching. This process was done without replacement, meaning that each individual was matched only once. To check whether the groups were well balanced after matching, we compared key characteristics, such as age, comorbidities, and vaccination status, using the standardized mean difference (SMD). An SMD less than 0.1 is generally considered to indicate that the groups are sufficiently similar for a meaningful comparison^32^.

### Mathematical model for SARS-CoV-2 viral dynamics and antiviral treatment

To describe the viral load trajectory over time, we used a state-of-the-art mechanistic model of viral dynamics^33,34^ (**Figure 1A**). The model tracks several key components of SARS-CoV-2 infection over time, including the number of susceptible cells (*S*), refractory cells (H), infected cells in the eclipse phase (*R*), productively infected cells (*I_E_*), and SARS-CoV-2 virions (*V*). Susceptible cells are infected at a rate proportional to the product of the infectivity rate, β, and the viral load, *V*. Once infected, cells transition to the eclipse phase, which a latent period lasting for a duration of 1/*k* on average before infected cells start to generate new virions at a production rate, π. Infected cells die due to viral cytopathic effects or are removed via the cytolytic innate immune response at a clearance rate, δ. While extended viral dynamic models for SARS-CoV-2 sometimes incorporate a delayed, time-varying adaptive immune response parameter33,34, we retained a parsimonious single-clearance structure. This choice prevents structural collinearity between the natural adaptive immune response and the antiviral treatment effect (which is typically initiated during the same temporal window), to ensure the robust and identifiable estimation of drug efficacy from the available clinical data. Free virions are removed from the system at a clearance rate γ. While not explicitly modelled, the model also incorporates the effect of immune responses where the production of interferons by infected cells induces susceptible cells into a temporary antiviral state. Here, susceptible cells are converted to a refractory state at a rate dependent on the number of productively infected cells (ϕ/*_P_*), while refractory cells can revert to a susceptible state at a rate *ρ*. The viral dynamics model (in the absence of antiviral treatment) can be described as a system of differential equations as follows:

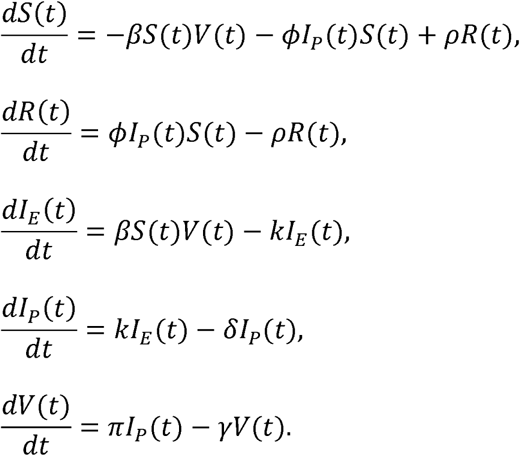

**Figure 1.**
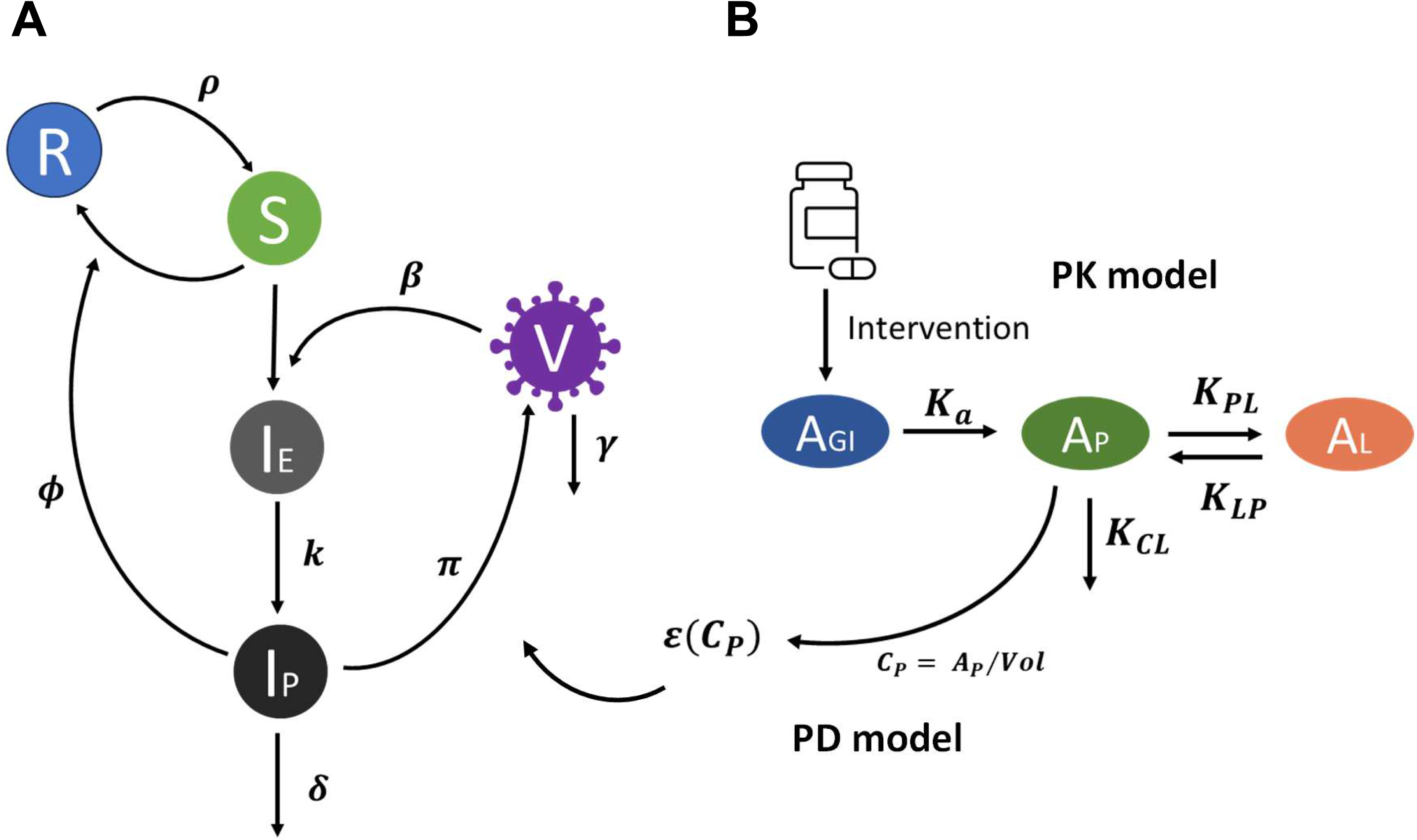
Schematic of the viral dynamics model and two-compartment PK-PD model. **(A)** The viral dynamics model tracks the following compartments: susceptible cells (*S*), refractory cells (*R*), eclipse-phase infected cells (*I_E_*), productively infected cells (*I_P_*), and free virus particles (*V*). The infection process occurs at rate β, with susceptible cells converting to refractory cells at rate φ and reverting back at rate ρ. Infected cells produce virus at rate π while is cleared at rate δ, and virus clearance rate is γ. **(B)** The two compartment PK-PD model describes the oral administration of nirmatrelvir and tracks the drug amount in the gut (*A*_GI_), plasma (*A*_P_), and tissue (*A*_L_). κ_a_ is the absorption rate of drug from gut to plasma, κ_PL_ and κ_LP_ are plasma-tissue transfer rate of the drug, κ_CL_ is the drug clearance rate. *C*_P_ is the drug concentration in plasma calculated by *C_P_* = *A_P_⁄Vol*, where *Vol* represents the estimated plasma volume. ε(*C*_P_) is the drug efficacy derived from the Hill equation, which blocks the viral reproduction in the viral dynamics model.

We modelled *v(t)* under the effect of antiviral treatment as follows:

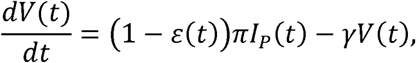

where *ε(t)* is the reduction of virus production due to the treatment at time *t*. We set the initial conditions of the viral dynamics model as {*S*= 10^7^, *R* = 0, *l_E_* = 0, *l_P_* = 0, *V* = 1}^35^ (**Supplementary Table 1**).

### Mathematical model for pharmacokinetics/pharmacodynamics

Following a previous study for the antiviral effect of nirmatrelvir-ritonavir^34^, we used a two-compartmental PK model with oral administration of nirmatrelvir–ritonavir (**Figure 1B**). The PK model is composed of three compartments: the gastrointestinal tract (*A*_GI_), plasma (*A*_P_), and tissue (*A*_L_). The drug is absorbed from the gastrointestinal tract at a constant rate, *κ_a_*, into the plasma compartment, from which the drug can be transferred to the lung tissue at a constant rate, *κ_PL_*, or be transferred back into the plasma compartment from the lung tissue at a constant rate, *κ_LP_*. The drug is subsequently removed from the body at a clearance rate, κ*_CL_*. At the start of the simulation (prior to any dosing), all compartments were initialized to zero, i.e., {*A_GI_* = 0,*A_P_* = 0,*A_L_* = 01. Oral dosing was implemented as an instantaneous addition to the gastrointestinal compartment. Specifically, at each dosing time *t= t_d_*, the amount in the GI compartment was updated according to Δ*A_GI_* = *Dose*. Following the treatment guidelines in China, we assumed a standard regimen of 300 mg nirmatrelvir boosted with 100 mg ritonavir, administered twice daily for five consecutive days. In this modeling framework, ritonavir was not explicitly modeled as a separate dynamic entity; instead, its pharmacokinetic boosting effect (inhibition of CYP3A4 metabolism) is intrinsically captured within the fixed pharmacokinetic parameters of nirmatrelvir (e.g., the reduced clearance rate, *κ_CL_*), which were derived from clinical data of the combination therapy^35^. Nirmatrelvir–ritonavir was administered twice daily, and repeat doses were therefore applied every 12 hours (e.g., *t*= 0, 12, 24, 36 h, etc.). Between dosing events, the system evolves according to the following PK equations:

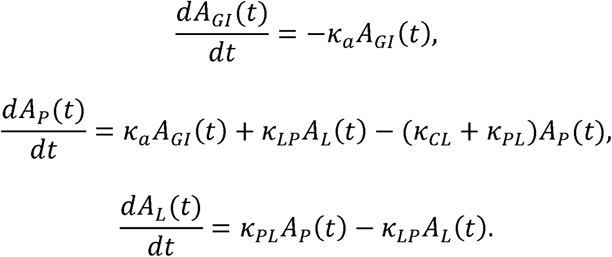

For the PD model, we assumed that the drug efficacy in blocking viral production is a function of the drug concentration in plasma, *C_P_* (i.e., amount of drug in plasma divided by the estimated plasma volume of distribution, *Vol*). Specifically, the drug efficacy, ε(*C_P_*), is estimated using the Hill equation^36^:

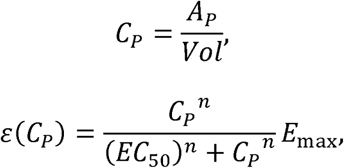

where *E*_max_ is the maximum efficacy, *n*, is the Hill coefficient, EC_50_ is the estimated in vivo effective concentration at which viral replication is reduced by 50%. We estimated the average treatment efficacy during the treatment as:

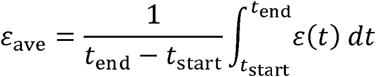

Given the absence of individual drug concentration or viral susceptibility measurements, treatment efficacy is modeled as an effective antiviral activity rather than a directly measurable pharmacodynamic quantity.

### Associations between patients’ characteristics and treatment efficacy

To evaluate the associations between patient demographic and clinical characteristics (e.g., age, sex, comorbidity status, and vaccination status) and antiviral treatment response, we employed a two-step post-hoc approach. First, individual Empirical Bayes Estimates (EBEs) for the viral dynamic and pharmacodynamic parameters (e.g., *EC*_50_) were extracted from the base structural model. Using these individual estimates and the simulated pharmacokinetic profiles, a time-averaged overall treatment efficacy (ε_ave_) over the treatment course was calculated for each patient using the aforementioned equations. Subsequently, multivariable linear regression was performed on these individual efficacy estimates to identify significant associations between patient characteristics and in vivo treatment response.

### Structural and statistical sensitivity analyses

To assess the robustness of the findings from our primary analysis, we performed sensitivity analyses with the following three different parameterizations.

#### i. Modelling with fixed upslope parameters β, π

Unlike high-frequency surveillance cohorts (such as the NBA cohort) that capture the early viral expansion phase, our retrospective RWD primarily consists of sparse, post-peak viral load measurements. Due to this lack of early-phase data, the viral infectivity β and virus production rates ε controlling the initial viral upslope could be practically nonidentifiable. While prior modeling efforts have resolved this unidentifiability by jointly modeling clinical trial data with the dense NBA cohort to inform early kinetics^34^, this data-pooling approach was not utilized for our matched RWD. To address this inherent data sparsity, we conducted a structural sensitivity analysis where β and π were fixed to the established Omicron-specific population priors (*log*₁₀β = 7.28; *log*₁₀ π = 2.72)^34^.

#### ii. Modelling without PKPD dynamics

To account for heterogeneities in PK parameters across individuals, we conducted a sensitivity analysis by removing the PK/PD components from the treatment model. Consequently, the viral load dynamics under treatment become:

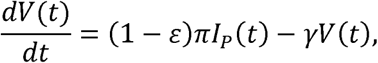

where ε is a treatment efficacy parameter (bounded between 0 and 1, following a logit-normal distribution), assumed to remain constant during the treatment window. To maintain symmetry with our primary analyses, this constant ε parameter was evaluated using both post-hoc extraction of EBEs for multivariable linear regression, and direct covariate estimation within the joint population model.

#### iii. Modelling with joint SAEM covariates

To rigorously verify that the covariate associations identified via linear regression were not statistical artifacts biased by EBE shrinkage, we conducted full nonlinear mixed-effects covariate analyses across our structural models. In this approach, patient characteristics were incorporated directly into the efficacy parameter (*EC*_50_) during the SAEM estimation step. This allowed us to evaluate the magnitude and significance of the covariate coefficients directly within the joint population model, independent of post-hoc EBE extraction.

### Parameter estimation: nonlinear mixed-effect modelling

We used a nonlinear mixed-effect approach to fit the viral dynamics model to the longitudinal viral load data. Fixed effects and random effects were estimated for each parameter using the following equations:

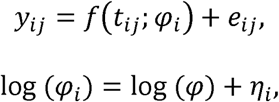

where *f* is the structural viral dynamics model, *Y_ij_* represents the observed viral load (log-transformed) for individual *i* at time *j*, and *ϕ_i_* denotes a vector of individual-specific viral dynamic parameters. Each individual’s parameter vector *ϕ_i_* was modelled as the sum of the population-level parameter vector φ and individual variability η*_i_*, which was assumed to follow a multivariate normal distribution: η*_i_*∼*N*(0,ω^2^).

## Results

### Patient characteristics

A total of 3,475 patients met the inclusion criteria (**Supplementary Figure 1**). **Table 1** summarizes the characteristics of the patients in the RWD before and after propensity score matching. Before matching, the control group was dominated by patients who were younger, male, had fewer comorbidities, and had higher vaccination coverage compared to the treatment group (**Table 1**). Furthermore, the viral load of the treatment group was consistently higher than that of the control group (**Figure 2, left**). After matching, the divergence in distributions between the control and treatment groups was reduced (SMDs < 0.1), with the exception of the age group (SMD = 0.1441) (**Table 1**). These results suggest that the matched control and treatment groups were comparable in our retrospective cohort. However, the viral load trajectories after matching were significantly different on Days 2, 4, 5, 6, 7, and 8 after detection (**Figure 2, right**), suggesting of an antiviral treatment effect, which we further investigate using the viral dynamics model.

**Table 1.** Characteristics of the SARS-CoV-2 patients in the real-world data from Shanghai, China.

| Variables |  | Before Matching |  | After Matching |  | Standardized<br>Mean<br>Difference<br>(SMD) |
| --- | --- | --- | --- | --- | --- | --- |
|  |  | Control<br>(N = 3246) | Treatment<br>(N = 229) | Control<br>(N = 229) | Treatment<br>(N = 229) |  |
| **Age** | 18-64 | 2291 (71%) | 49 (21%) | 82 (36%) | 49 (21%) | - |
|  | ≥ 65 | 955 (29%) | 180 (79%) | 147 (64%) | 180 (79%) | 0.1441 |
| **Sex** | Female | 1398 (43%) | 116 (51%) | 132 (58%) | 116 (51%) | - |
|  | Male | 1848 (57%) | 113 (49%) | 97 (42%) | 113 (49%) | 0.0699 |
| **Number of**<br>**comorbidities** | 0 | 1697 (52%) | 57 (25%) | 61 (27%) | 57 (25%) | - |
|  | ≥ 1 | 1549 (48%) | 172 (75%) | 168 (73%) | 172 (75%) | 0.0175 |
| **Vaccination**<br>**status** | < 2 doses | 1694 (52%) | 145 (63%) | 132 (58%) | 145 (63%) | 0.0568 |
|  | 2 doses | 678 (21%) | 47 (21%) | 55 (24%) | 47 (21%) | -0.0349 |
|  | 3 doses | 874 (27%) | 37 (16%) | 42 (18%) | 37 (16%) | 0.0218 |
| **Initial viral load** | -- | 5.15 (1.97) | 7.12 (1.86) | 7.15 (1.76) | 7.12 (1.86) | -0.0165 |

**Figure 2.**
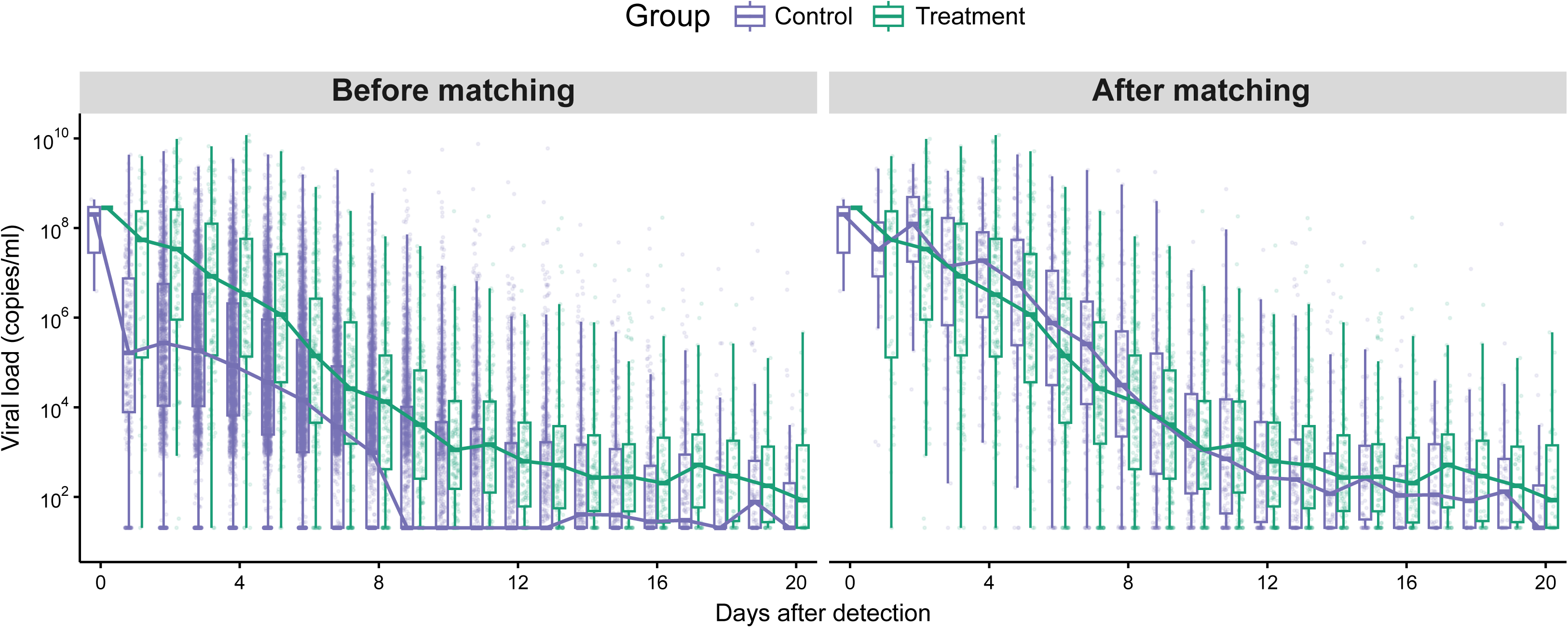
Longitudinal comparison of SARS-CoV-2 viral load trajectories between treatment and control groups. Boxplots show the distribution of viral load (log_10_ copies/mL) over time from detection (Day 0 to Day 16) for individuals in the treatment group (green) and the control group (purple) before propensity score matching (left) and after propensity score matching (right). Each point represents an individual measurement. The median, interquartile range, and overall spread are displayed for each group at each time point. Medians at each day were connected by lines. Asterisks correspond to the time points where the viral load distributions were statistically significantly different between the control and treatment groups (*p* < 0.05, t-test).

### Effect of nirmatrelvir–ritonavir on reducing viral replication

We fitted the mathematical model to quantify the viral load dynamics with and without nirmatrelvir–ritonavir treatment. We estimated that nirmatrelvir–ritonavir treatment reduced the virus production by 55.2% over the 5-day treatment period under the assumed PK–PD structure. All estimated parameters for the viral dynamics and PK/PD models are reported in **Supplementary Table 1**.

**Figure 3** presents modelled trajectories of viral load and drug concentration under different treatment initiation timings, simulated using the population parameter estimates. We observed a viral rebound, defined as a resurgence of viral load after initial improvement shortly after treatment, depending on the timing of treatment initiation. When treatment is administered shortly after infection (on Day 0 of detection) and before the viral load peak, the reduction in peak viral load is minimal, and a notable rebound is observed following treatment cessation (**Figure 3AB, left**). On the other hand, if treatment begins after the peak of viral load, the viral load declines during the treatment period and reaches the detection limit earlier than in untreated cases, with no observable rebound (**Figure 3AB, right)**. **Supplementary Figures 2 and 3** show the estimated viral load curve for each individual.

**Figure 3.**
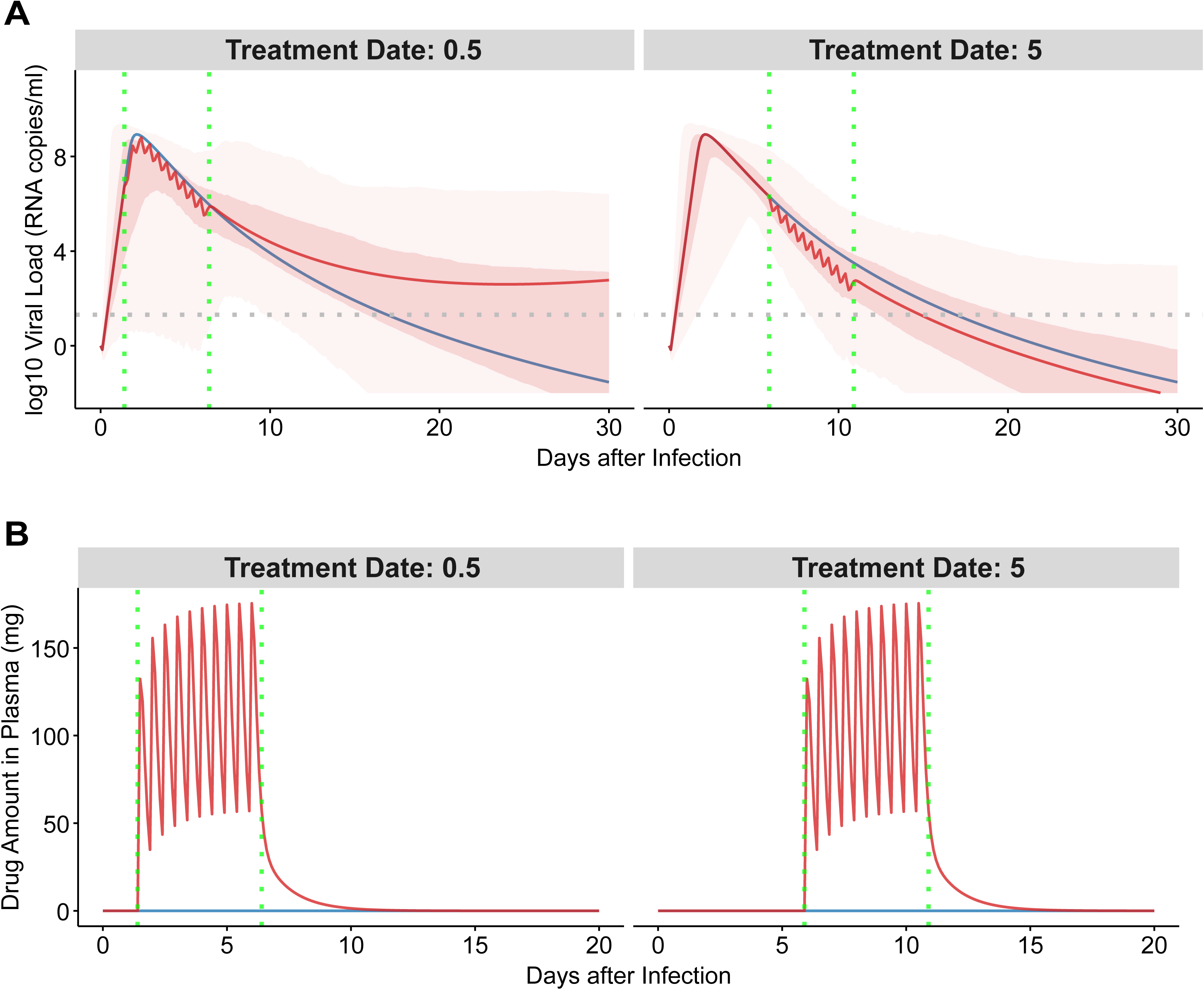
Modeled viral load trajectories and drug concentration in plasma stratified by timing of treatment initiation. **(A)** Predicted upper respiratory tract viral load dynamics (log_10_ RNA copies/mL) are shown for individuals grouped by treatment start time: Day 0 (left), Day 5 (right) after the day of the first detection. Solid lines represent the best-fitting model using the population parameters (red for treated group, blue for untreated group), and shaded areas denote the 50% and 95% confidence intervals of treated group. The horizontal dashed line represents the assay’s detection limit (10^1.31^ copies/mL). **(B)** Predicted nirmatrelvir concentration in plasma for individuals (red for treated group, blue for untreated group) stratified by treatment start time: Day 0 (left), Day 5 (right) after the day of the first detection. In both panels, vertical dotted lines indicate treatment initiation (first line) and 5 days post-treatment start (second line), corresponding to a typical antiviral treatment course in China.

### Model-derived viral shedding duration across age, vaccination, and treatment groups

Using the individual-level viral load trajectories simulated from the best-fitting viral dynamics and PK/PD model, we calculated each patient’s viral shedding duration, defined as the time interval between the first simulated viral load measurement exceeding the assay’s detection threshold (1.31 log__ copies/mL) and the first subsequent time point at which the simulated viral load dropped below this threshold (**Figure 4**). For individuals whose simulated viral load remained above 1.31 throughout the entire 40-day simulation window, the shedding duration was set to 40 days. **Figure 4** shows the distribution of model-derived shedding durations stratified by treatment status, age group, and vaccination status. We found that the impact of nirmatrelvir–ritonavir on shortening shedding duration varied markedly across subgroups. Among younger adults (18–64 years), treatment significantly reduced shedding only in individuals who had received three vaccine doses (p ≈ 0.009). In contrast, no statistically significant reduction in shedding duration was observed among younger adults with fewer than three doses, or among older adults (≥65 years) across all vaccination strata (all p > 0.20). These findings suggest that the effect of nirmatrelvir–ritonavir on viral clearance varies across age and vaccination status.

**Figure 4.**
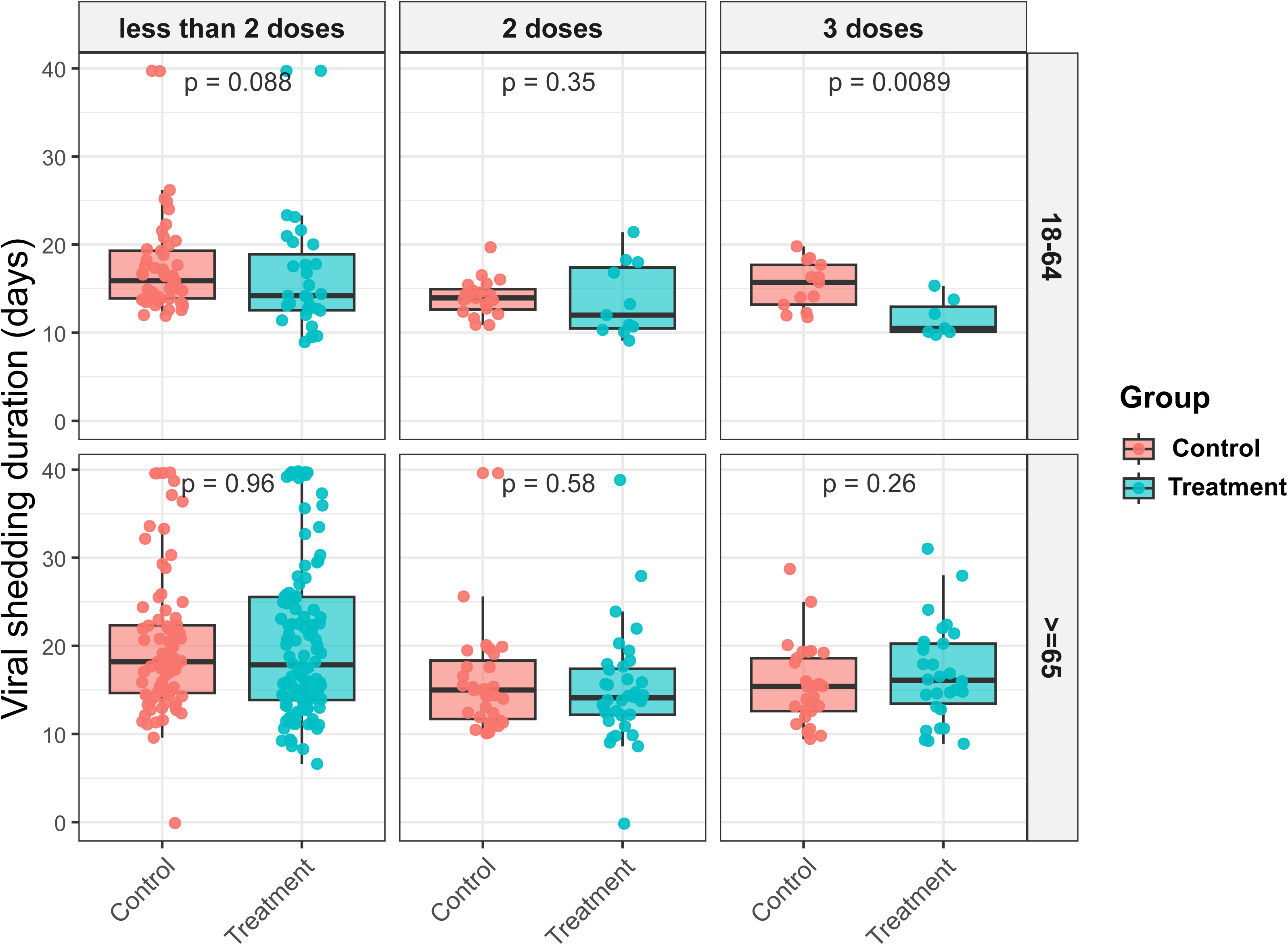
Model-derived viral shedding duration stratified by age, vaccination status, and treatment. Shedding duration was computed from each individual’s simulated viral load trajectory as the time from the first model-predicted viral load above the detection threshold (1.31 log__ copies/mL) to the first subsequent time point below this threshold. For individuals whose viral load remained above 1.31 throughout the 40-day simulation, shedding duration was set to 40 days. Boxplots and jittered points show shedding duration across treatment groups and subgroups defined by age (18–64 vs ≥65 years) and vaccination (<2, 2, or 3 doses). p-values indicate Wilcoxon rank-sum tests comparing treated and untreated individuals within each subgroup.

### Association between viral dynamics, treatment efficacy, and patient characteristics

We observed substantial inter-individual heterogeneity in the posterior distributions of both viral dynamics and treatment response parameters. **Supplementary Figure 4** shows the distribution of the parameters, which were estimated for each individual patient. The individual parameters of viral dynamics, such as the viral infectivity rate (β), viral production rate (π), refractory to susceptible reversion rate (ρ), and infected cell clearance rate (δ), varied widely across individuals (**Supplementary Figures 4A-E**). Furthermore, for the individual parameters of the PK/PD model, the in vivo effective concentration (*EC_50_*)also showed substantial dispersion, suggesting large heterogeneity in treatment response (**Supplementary Figure 4F**).

To understand the source of such heterogeneity in the model parameters, we performed a multivariable linear regression analysis using the individual parameter values as dependent variables and the patient characteristics (i.e., age, sex, comorbidity, vaccination status) as independent categorical variables. Younger age (< 65 years) and more vaccination doses were associated with faster infected cell clearance, whereas older age and less vaccination doses were linked to prolonged viral shedding, consistent with our previous findings^15^ (**Table 2**). Treatment efficacy also varied by age and vaccination status. Specifically, older adults (≥ 65 years) had 7% (95% CI: [1%, 13%]) lower treatment efficacy compared to younger adults (18–64 years). In contrast, individuals who received 2 or 3 doses of vaccination showed 8% (95% CI: [2%, 14%]) and 12% (95% CI: [5%, 18]) higher antiviral efficacy, respectively, compared to those who received less than 2 doses of vaccination (**Table 2**). The associations with comorbidity and sex were not statistically significant. Figure 5 shows the association between age/vaccination status and treatment efficacy, including both the raw data and the model prediction. **Supplementary** Figure 5 shows the individual treatment efficacy stratified by age and vaccination status. **Supplementary** Figure 6 shows the modelled individual viral load trajectories under treatment, stratified by vaccination status and age, highlighting a more rapid decline in viral load in patient that received more doses of vaccination and in younger individuals.

**Figure 5.**
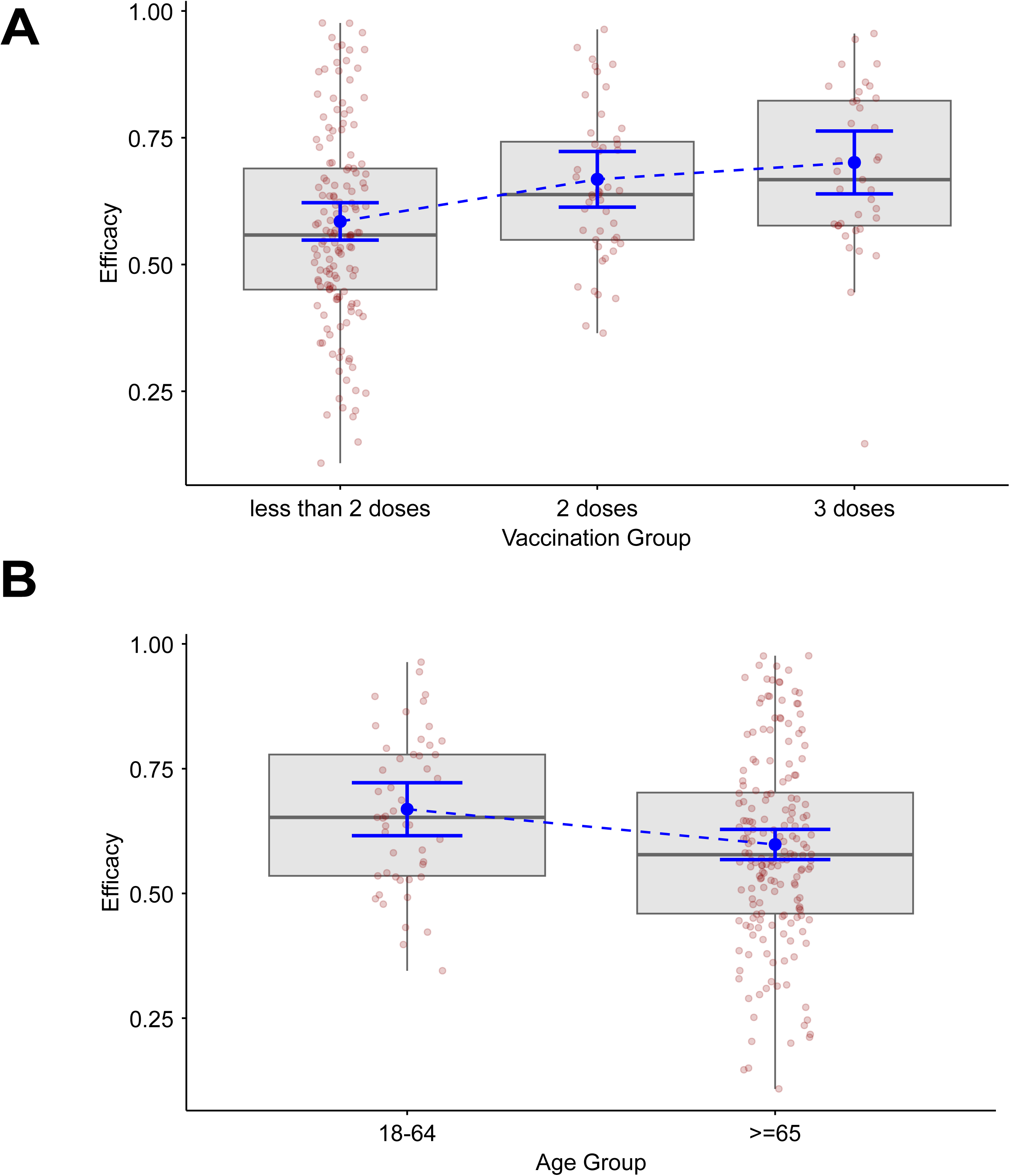
Associations between key clinical factors (age, vaccination) and treatment efficacy. **(A)** Model-estimated treatment efficacy across vaccination groups (<2 doses, 2 doses, and 3 doses). **(B)** Model-estimated efficacy by age group (18–64 vs ≥65 years). Brown points represent individual efficacy values, boxplots show the distribution of raw data. The blue points with error bars represent predicted means and 95% confidence intervals derived from a multivariable linear regression model adjusted for categorized age, vaccination status, comorbidity, and sex. The blue dashed lines connecting model prediction means are only for visualization purpose instead of implying continuity.

**Table 2.** Association between SARS-CoV-2 patient characteristics and the model parameters^a^.

|  |  | Viral Dynamics Model |  |  |  |  | PK/PD Model |  |  |
| --- | --- | --- | --- | --- | --- | --- | --- | --- | --- |
|  |  | Viral infectivity<br>(log<sub>10</sub>(RNAcopies/mL)day<sup>-1</sup>) | Viral<br>production<br>rate (log<sub>10</sub>day<sup>-1</sup>) | Refractory<br>to<br>susceptible<br>reversion<br>rate (log<sub>10</sub>day<sup>-1</sup>) | Susceptible<br>to refractory<br>conversion<br>rate (log<sub>10</sub>cell<sup>-1</sup><br>day<sup>-1</sup>) | Infected cell<br>clearance<br>rate (day<sup>-1</sup>) | Onset of acquired<br>immunity relative<br>to detection (days) | In-vivo effective<br>concentration<br>EC<sub>50</sub> | Average<br>treatment<br>efficacy |
| Symbol |  | log<sub>10</sub>β | log<sub>10</sub>π | log<sub>10</sub>ρ | log<sub>10</sub>φ | δ | τ | EC<sub>50</sub> | ε<sub>ave</sub> |
| Age | 18-64 | Reference |  |  |  |  |  |  |  |
|  | ≥ 65 | -0.02<br>(-0.10, 0.05) | 0.00<br>(-0.04, 0.03) | **0.05\***<br>**(0.00, 0.09)** | 0.00<br>(0.00, 0.01) | **-0.08\***<br>**(-0.15, -0.01)** | -0.10<br>(-0.60, 0.41) | **0.65 \***<br>**(0.13, 1.18)** | **-0.07\***<br>**(-0.13, -0.01)** |
| Sex | Female | Reference |  |  |  |  |  |  |  |
|  | Male | 0.03<br>(-0.04, 0.10) | -0.01<br>(-0.04, 0.02) | 0.01<br>(-0.03, 0.05) | 0.00<br>(0.00, 0.00) | -0.01<br>(-0.06, 0.05) | 0.20<br>(-0.22, 0.61) | 0.28<br>(-0.15, 0.71) | -0.02<br>(-0.07, 0.03) |
| Number of<br>comorbidities<sup>*</sup> | 0 | Reference |  |  |  |  |  |  |  |
|  | ≥1 | -0.05<br>(-0.12, 0.03) | 0.00<br>(-0.04, 0.03) | 0.01<br>(-0.03, 0.06) | 0.00<br>(0.00, 0.00) | 0.00<br>(-0.06, 0.07) | -0.04<br>(-0.51, 0.44) | -0.24<br>(-0.73, 0.26) | 0.02<br>(-0.03, 0.08) |
| Vaccination<br>status | < 2 doses | Reference |  |  |  |  |  |  |  |
|  | 2 doses | 0.02<br>(-0.06, 0.10) | -0.01<br>(-0.05, 0.03) | **-0.05\***<br>**(-0.10, 0.00)** | 0.00<br>(-0.01, 0.00) | **0.10\*\***<br>**(0.03, 0.17)** | 0.02<br>(-0.51, 0.54) | **-0.79 \***<br>**(-1.33, -0.25)** | **0.08\***<br>**(0.02, 0.14)** |
|  | 3 doses | 0.02<br>(-0.07, 0.11) | -0.02<br>(-0.06, 0.03) | -0.04<br>(-0.09, 0.01) | 0.00<br>(-0.01, 0.00) | **0.10\***<br>**(0.02, 0.18)** | 0.03<br>(-0.54, 0.60) | **-1.00\*\***<br>**(-1.59, -0.40)** | **0.12\*\***<br>**(0.05, 0.18)** |

<sup>a</sup>the numbers in parentheses are 95% confidence intervals
\* $p < 0.05$ ; \*\* $p < 0.01$ ; \*\*\* $p < 0.001$

### Sensitivity analyses: Association between patients’ age & vaccination status and treatment efficacy

To evaluate the robustness of these primary findings against alternative structural and statistical assumptions, a comprehensive series of sensitivity analyses was conducted (**Supplementary Tables S2 and S3**). When patient characteristics were evaluated jointly within the population model to account for potential shrinkage bias (M5), the significant associations of both age (Age > 65, β = 0.42; p<0.05) and vaccination (Doses ≥ 2, β = -0.59; p<0.001) with EC_50_ were confirmed. These associations also remained consistent when the time-varying PK/PD components were simplified to a constant efficacy (ε) parameter (M3, M7).

However, divergent results were observed in the structurally constrained sensitivity models where early-phase viral infectivity and production rates were fixed and individual variability was removed (M2, M4, M6, M8). In these constrained models, the association between older age and reduced treatment efficacy lost formal statistical significance across both the post-hoc and joint estimation frameworks. Notably, higher vaccination doses remained a consistently significant association of enhanced treatment efficacy across all evaluated structural parameterizations and statistical methodologies. Together, these sensitivity analyses demonstrate that the positive effect of vaccination on in vivo antiviral efficacy is highly robust, whereas the initial age-related associations are sensitive to the unobserved variations in baseline early viral kinetics.

## Discussion

We investigated whether patient characteristics influence the antiviral efficacy of nirmatrelvir–ritonavir using real-world data from patients infected with the SARS-CoV-2 BA.2 variant in China. Our findings suggest that the effectiveness of nirmatrelvir–ritonavir in reducing viral load is associated with both age and vaccination status, with greater efficacy observed in younger individuals and those who have received two or more vaccine doses. These results have important clinical implications, highlighting the potential for more personalized antiviral treatment strategies, such as optimizing treatment duration and dosing regimens.

We observed that early administration of nirmatrelvir–ritonavir can lead to a rebound in viral load following the standard 5-day treatment period. This phenomenon has been previously documented in both clinical and modelling studies. For instance, a clinical cohort study by Pandit et al. reported that 14.2% of individuals treated with nirmatrelvir–ritonavir experienced viral rebound^37^. In addition, Esmaeili et al. elucidated the underlying mechanism through a mathematical modelling approach, and found that extending the duration of treatment, rather than increasing the dose or dosing frequency, may be more effective in preventing viral rebound^34^.

RNA viruses have been found to have enhanced replication efficacy in senescent cells that accumulate with age, which may imply that the viral replicative machinery, including RdRp and M^pro^, are more active in older adults infected with SARS-CoV-2^38–40^. Moreover, the angiotensin-converting enzyme 2 (ACE2), which SARS-CoV-2 S glycoprotein directly interacts with as an entry point into cells, is also upregulated in the senescent phenotype^41^, thereby further increasing the rates of viral entry and replication. Additionally, IL-6, a key marker of cell aging and an inflammatory cytokine overexpressed in the frailty-related hyperinflammatory state, has been associated with higher SARS-CoV-2 viral load^42^, possibly related to dysregulated immune responses which are less able to counteract viral replication^43^. Indeed, a recent study by Mafi et al. reported that nirmatrelvir–ritonavir was not effective in reducing the risk of hospitalization or all-cause mortality in the elderly population^44^. Furthermore, Lin et al. found that the clinical efficacy of nirmatrelvir-ritonavir in immunocompromised individuals was inferior as compared to immunocompetent patients^45^. As such, the overall increase in viral replicative capacity — due to immunosenescence, lack of immunity conferred by vaccination, or an immunocompromised state — may diminish the relative efficacy of M^pro^ inhibitors on reducing the viral load, even when the pharmacological potency remains constant.

A finding of our study bridges these biological observations with our model estimates. In our primary unconstrained analysis, older age was associated with reduced antiviral potency. However, this association was diminished in our structural sensitivity analyses when early-phase viral kinetic rates (β, π) were fixed to biological priors. This discrepancy highlights a known identifiability challenge when modeling sparse, post-peak real-world data. Without early viral load measurements to reliably anchor the initial viral expansion, the unconstrained model absorbed the unobserved variance in baseline kinetics—such as the higher viral replication driven by senescence and immune dysregulation in older adults—into the treatment efficacy parameter. The absence of the age effect in our constrained models strongly suggests that the intrinsic pharmacological potency of the antiviral agent is maintained in older adults, but their overall treatment response is masked by a much tougher natural baseline disease trajectory. Conversely, vaccination remained a highly significant predictor of enhanced treatment efficacy across every structural and statistical sensitivity test, confirming that prior immunity provides a true, independent enhancement to the net antiviral blockade.

This is not the first study to investigate the efficacy of nirmatrelvir–ritonavir in reducing viral load using within-host modelling approaches. Esmaeili et al. conducted a similar analysis using data from nirmatrelvir–ritonavir clinical trials^34^, and also reported that viral rebound may be induced by early treatment and that the in vivo potency of nirmatrelvir–ritonavir is substantially lower than what is observed in in vitro assays. They found significant differences in potency between two trials: the EPIC-HR trial^46^, which involved high-risk adults (i.e., patients with one or more of the following risk factors: age > 60 years, BMI > 25, smoking, immunosuppressive disease, or chronic lung, cardiovascular, kidney, or cancer diseases), showed lower potency compared to the PLATCOV trial^47^, which enrolled lower-risk adults without comorbidities. For example, the authors estimated an average treatment efficacy in the EPIC-HR trial of 82%. On the other hand, our estimate based on our retrospective cohort was substantially lower (55.2%), further supporting the importance of using similar cohort to compare estimates of antiviral treatment. It is important to note that the absolute magnitude of estimated antiviral efficacy is inherently model-dependent and sensitive to assumptions about the pharmacodynamic concentration–response relationship. In the absence of individual pharmacokinetic or in vivo susceptibility measurements, different modeling choices can lead to substantially different numerical efficacy estimates, even under similar drug concentration profiles. For this reason, we focus on relative differences in effective antiviral activity across patient subgroups and assess the robustness of these associations through sensitivity analyses. Moreover, differently from Esmaeili et al.^34^, our study help shedding light on the underlying population characteristics that are the source of the observed heterogeneities in treatment effect.

This study has several strengths. First, we showed the potential impact of aging and vaccination status on antiviral treatment efficacy using RWD with a mechanistic modelling approach. The incorporation of RWD in the analysis reflects the broader effectiveness and practical use of interventions in routine clinical settings, thus contributing unique insights into treatment efficacy. Second, we quantified the antiviral effect on viral load dynamics using a mechanistic model, which enables not only estimation of treatment impact but also exploration of underlying biological processes, such as age- and vaccine-dependent differences in drug metabolism or immune response. This mechanistic insight goes beyond empirical associations and contributes to a more mechanistically informed framework for treatment evaluation. Our modeling indicates that older adults (over 65 years) may be effectively under-treated by the standard protocol. Because older adults exhibit slower natural viral clearance (lower δ) combined with higher baseline viral replication trajectories, the standard 5-day course frequently terminates before their immune system can secure viral elimination. This suggests that ’personalized treatment’ for the elderly should not just mean prioritizing them for treatment access, but optimizing treatment regimen for the elderly, such as extending their treatment course to prevent the viral rebound. Third, our mathematical model-based analyses provided forward-looking insights into potential outcomes under various hypothetical conditions (e.g., earlier treatment initiation, different age groups, or varying levels of vaccination coverage). This approach supports scenario planning and individualized treatment strategies, especially for high-risk populations.

A set of limitations should be acknowledged as well. First, we only considered a proxy for viral load that is obtained by the analysis of RT-qPCR data from the upper respiratory tract (nasopharyngeal or oropharyngeal samples). Studying other sample types may provide more nuanced information on the effect of antivirals on viral load reduction. For instance, a remdesivir trial showed that while no significant decrease in upper respiratory tract viral load between the treatment and control groups; however, it was effective in significantly reducing SARS-CoV-2 viremia^48^. Second, we fixed the PK parameters for nirmatrelvir–ritonavir, which might also be influenced by patient characteristics. However, we performed a sensitivity analysis without PK/PD dynamics and consistently found that age and vaccination status modify treatment effect. It is important to note that each participant’s estimated treatment efficacy (ε) is effectively a composite function of both pharmacokinetic (PK) and pharmacodynamic (PD) variability. In this study, we did not measure individual-level drug concentrations or in vivo viral susceptibility (e.g., EC__), making it impossible to disentangle the relative contributions of PK versus PD variability to the observed heterogeneity in treatment response. This suggests the possibility for future studies to incorporate individualized drug level measurements and viral phenotyping to better understand the mechanistic basis of differential antiviral efficacy. Third, we only considered nirmatrelvir–ritonavir in the RWD analyses due to the limited data for other antivirals such as remdesivir and molnupiravir. The analysis could be extended to other antivirals to better understand and inform the standard of care practice for COVID-19 patients. Fourth, the data is limited to patients infected with the Omicron BA.2 variant. Estimates considering patients infected with other SARS-CoV-2 variants may provide further support and/or extend our findings. Fifth, our real-world dataset inherently lacks high-frequency viral load sampling during the early, pre-peak viral expansion phase. Because early-phase infectivity and viral production parameters are collinear with treatment efficacy in structural models, this data sparsity could lead to parameter unidentifiability. While we robustly addressed this through fixed-kinetic sensitivity modeling—revealing that age-related declines in efficacy could be artifacts of unobserved baseline viral dynamics, future prospective studies with dense early swabbing are required to fully disentangle initial viral expansion rates from subsequent antiviral drug action. Lastly, although clinical outcomes such as disease severity, hospitalization, or mortality are commonly used measures of antiviral effect, their analysis in this real-world dataset is complicated by indication bias. In routine clinical practice, nirmatrelvir–ritonavir is preferentially prescribed to older individuals and patients with more severe disease at presentation (**Table 1**), making post-treatment severity an unsuitable outcome for comparison without fair definition of clinical improvement/deterioration. For this reason, we focused on mechanistic effect of antivirals (i.e., reduction in virus production), which represents a proximal and interpretable marker of antiviral activity and are less directly influenced by treatment allocation decisions.

In summary, our results indicate that patient-specific factors—most robustly vaccination status, alongside age-related viral kinetics—significantly dictate the treatment effect of nirmatrelvir–ritonavir. The observed heterogeneity likely reflects a complex interplay between true pharmacological enhancement from prior immunity and differing natural virological trajectories among individuals. By incorporating these variables, our modeling approach yields mechanistic insights into treatment responses and helps lay the groundwork for designing personalized antiviral treatment strategies.

## Data Availability

The data that support the findings of this study are not publicly available due to data governance and privacy restrictions but are available from the corresponding authors upon reasonable request and with permission from the data providers.

## Data Availability

The data supporting the findings of this study are available from the corresponding authors on request.

## Code Availability

All analyses were performed with the statistical computing software R (version 4.3.3). The analysis using nonlinear mixed effects models was performed on MONOLIX 2019R2 (www.lixoft.com). The analytical codes for this study can be found at the following repository: https://github.com/YiyuL121/Shanghai-RWD.

## Acknowledgments

This study was supported in part by the Ministry of Education, Singapore, under the Academic Research Fund Tier 1 Seed Award (RLMOE100201900000001) (to KE), a Singapore Ministry of Education startup grant (LKCMedicine-SUG, #022487-00001) (to KE), JST, PRESTO (JPMJPR23R3) (to KE).

## Author Contributions

Conceived and designed the study: WZ, HY, KE. Obtained and analyzed the data: LY, Yan Wang, Yuqian Wang, JA, LBK, DL, JZ, HW, Yanpeng Wu, KE. Wrote the paper: LY, Yan Wang, Yuqian Wang, DL, WZ, HY, KE. Edited the paper: LY, Yan Wang, Yuqian Wang, LBK, DL, PYC, HKC, CEZC, JTS, KO, SE, BJC, MEC, HS, LEW, BEY, MA, TMN, ECYC, WZ, HY, KE. All authors read and approved the final manuscript.

## Supplementary Information

**Supplementary Figure 1. Patient selection flowchart for inclusion in the analysis.** Of 48,243 patients initially identified, 44,768 were excluded based on predefined criteria, resulting in 3,475 patients remaining in the final analysis. Note on treatment allocation: During the outbreak, adequate drug stockpiles allowed for treatment administration based on clinical practice considerations. Specifically, 81 participants without high-risk factors received nirmatrelvir–ritonavir upon strong request to alleviate anxiety in isolation wards, after excluding contraindications. Conversely, some patients with high-risk factors did not receive treatment due to contraindications such as impaired hepatic or renal function, or drug-drug interactions. To mitigate selection bias, our final analysis was strictly restricted to patients meeting the defined risk criteria.

**Supplementary Figure 2. Individual-level viral load trajectories with model fits and uncertainty bounds for the untreated patients.** Each panel represents the observed and modeled viral load dynamics for a single patient (not treated by any antivirals) over time since infection. Blue dots indicate observed viral load data, and red lines show the best-fitting model for each individual. The shaded pink area represents the 95% confidence interval of the model fit. The horizontal dashed line represents the assay’s detection limit (10^1.31^ copies/mL).

**Supplementary Figure 3. Individual-level viral load trajectories with model fits and uncertainty bounds for the treated patients.** Each panel represents the observed and modeled viral load dynamics for a single patient (treated with nirmatrelvir–ritonavir) over time since infection. Blue dots indicate observed viral load data, and red lines show the best-fitting model for each individual. The shaded pink area represents the 95% confidence intervals of the model fit. The horizontal dashed line represents the assay’s detection limit (10^1.31^ copies/mL). Green dashed vertical lines indicate treatment initiation dates.

**Supplementary Figure 4. Distributions of individual-level parameter estimates from the viral dynamics and PKPD models.** Parameter values estimated for each individual are shown as histograms. The estimated parameters of the viral dynamics models include the (**A**) viral infectivity rate (log /3), (**B**) viral production rate (log n), (**C**) refractory to susceptible reversion rate (log p), (**D**) susceptible to refractory conversion rate (log ¢), and (**E**) infected cell clearance rate (δ). The estimated parameter of the PK/PD model includes the (**F**) potency reduction factor (prf).

**Supplementary Figure 6. Demonstration of viral load trajectories under treatment across different patients’ characteristics.** The trajectories and confidential intervals are computed based on estimated individual viral load values (reported in S-Fig-3) of specific groups, without considering other factors. The solid line is the numeric mean, and shaded area represents the 95% quantile of the individual viral load values within one group. (**A**) The treated group is stratified by vaccination status. (**B**) the treated group is stratified by the age of 65 years old.

**Supplementary Table 1.** Population parameter values for the viral dynamics and PK/PD models.

|  |  |  |  |  |  |  |
| --- | --- | --- | --- | --- | --- | --- |
| **Parameter (unit)** | **Symbol** | **Population Mean** | **Standard Error** | **Standard Deviation of Random Effects** | **Distribution** | **Source** |
| **Viral dynamics model** |  |  |  |  |  |  |
| Viral infectivity<br>(log<sub>10</sub> (RNA copies/mL) day<sup>-1</sup>) | $\log_{10}\beta$ | -7.24 | 0.11 | 0.42 | Normal | Estimated |
| Viral production rate<br>(log<sub>10</sub> day<sup>-1</sup>) | $\log_{10}\pi$ | 3.43 | 4.25e-2 | 0.29 | Normal | Estimated |
| Refractory to susceptible reversion rate (log<sub>10</sub> day<sup>-1</sup>) | $\log_{10}\rho$ | -1.12 | 4.95e-2 | 0.35 | Normal | Estimated |
| Susceptible to refractory conversion rate (log<sub>10</sub> cell<sup>-1</sup> day<sup>-1</sup>) | $\log_{10}\phi$ | -6.93 | 1.6e-2 | 0.061 | Normal | Estimated |
| Infected cell clearance rate (day<sup>-1</sup>) | $\delta$ | 1.83 | 3.7e-2 | 0.19 | Lognormal | Estimated |
| Delay between infection and detection (days) | $\tau$ | 0.93 | 0.21 | 1.19 | Logit[0,20] | Estimated |
| Initial viral inoculum (RNA copies/mL) | $V(0)$ | 1 | -- | -- | -- | Fixed |
| Viral clearance rate (day<sup>-1</sup>) | $\gamma$ | 15 | -- | -- | -- | 49 |
| Mean eclipse phase duration (days<sup>-1</sup>) | $1/k$ | 1/4 | -- | -- | -- | 50 |
| Initial number of susceptible cells | $S(0)$ | 1e7 | -- | -- | -- | 49 |
| Initial number of refractory cells | $R(0)$ | 0 | -- | -- | -- | -- |
| Initial number of productively infected cells | $I(0)$ | 0 | -- | -- | -- | -- |
| Initial number of infected cells in eclipse phase | $I(0)$ | 0 | -- | -- | -- | -- |
| **PK/PD model** |  |  |  |  |  |  |
| Absorption rate constant (day<sup>-1</sup>) | $\kappa$ | 9.98 | -- | -- | -- | 34 |
| Rate constant of transfer from plasma to lung (day<sup>-1</sup>) | $\kappa_{PL}$ | 1.58 | -- | -- | -- | 34 |
| Rate constant of transfer from lung to plasma (day<sup>-1</sup>) | $\kappa_{LP}$ | 1.22 | -- | -- | -- | 34 |
| Elimination rate constant (day<sup>-1</sup>) | $\kappa_{CL}$ | 4.96 | -- | -- | -- | 34 |
| Plasma volume of distribution (mL) | $Vol$ | 41,743 | -- | -- | -- | 34 |
| Maximum efficacy (%) | $E_{\max}$ | 99.9 | -- | -- | -- | 34 |
| In vivo effective concentration (μM) | $EC_{50}$ | 4.35 | 0.54 | 0.81 | Lognormal | Estimated |
| Hill coefficient | $n$ | 3.16 | -- | -- | -- | 34 |
| Potency reduction factor | prf | 49.4 | -- | -- | -- | Computed<sup>34</sup> |
| Average treatment efficacy (%) | $\varepsilon_{ave}$ | 55.24 | -- | -- | -- | Computed |

**Supplementary Table S2.** Sensitivity analysis comparing alternative model parameterizations and covariate effects.

|  |  |  |  |  |  |
| --- | --- | --- | --- | --- | --- |
| Model Specifications |  |  |  | Akaike Information Criteria (AIC) | Significant Association(s) with Efficacy <sup>d</sup> |
| Type | Definition | ID | Parameterization <sup>a,b,c</sup> |  |  |
| Structural base models <sup>a</sup> | TREIV model with structural variations | M1 | Unconstrained Base model (Estimate $\beta$, $\pi$, $\rho$, □, $\delta$, $\tau$, $EC_{50}$) | 16188 | Age, Vaccinations |
| | | M2 | Fixed kinetic model ($\beta_{pop}$, $\pi_{pop}$ fixed) | 16283 | Vaccinations |
| Modifying PKPD component <sup>b</sup> | TREIV model with PKPD variations | M3 | Constant efficacy $\varepsilon$ adjusted from M1 | 16205 | Age, Vaccinations |
| | | M4 | Constant efficacy $\varepsilon$ adjusted from M2 | 16402 | Vaccinations |
| Covariate models <sup>c</sup> | TREIV model with defined covariates | M5 | Age, Vaccine as covariates on $EC_{50}$ adjusted from M1 | 16172 | Age, Vaccinations |
| | | M6 | Age, Vaccine as covariates on $EC_{50}$ adjusted from M2 | 16367 | Vaccinations |
| | | M7 | Age, Vaccine as covariates on $\varepsilon$ adjusted from M3 | 16180 | Age, Vaccinations |
| | | M8 | Age, Vaccine as covariates on $\varepsilon$ adjusted from M4 | 16396 | Vaccinations |

<sup>a</sup> M1: Primary base model; M2: Primary base model with fixed upslope parameters.
<sup>b</sup> Models without PKPD components, efficacy was estimated.
<sup>c</sup> Covariate models where categorized age and categorized vaccination status serve as covariates on $EC_{50}$ or $\varepsilon$
<sup>d</sup> For models M1, M2, M3, M4: Associations were determined by regression on EBEs. For models M5, M6, M7, M8: Associations were determined by Wald Test on covariate effects.

**Supplementary Table S3.** Wald test results for evaluated covariates on treatment efficacy across alternative population models: M5-M8. ^a,b,c^.

|  |  |  |  |
| --- | --- | --- | --- |
| Model ID <sup>a</sup> | Model Description | Covariates on EC<sub>50</sub>/ $\varepsilon$ | Estimate ($\beta$) <sup>b, c</sup> |
| M5 | Primary Model M1 with covariates on EC<sub>50</sub> | Age (> 65 years) | 0.42* |
| | | Vaccination (≥ 2 doses) | -0.59*** |
| M6 | Fixed kinetic model M2 with covariates on EC<sub>50</sub> | Age (> 65 years) | 0.27 |
| | | Vaccination (≥ 2 doses) | -0.69*** |
| M7 | Constant efficacy model M3 with covariates on $\varepsilon$ | Age (> 65 years) | -1.22** |
| | | Vaccination (≥ 2 doses) | 1.34*** |
| M8 | Fixed kinetic & Constant efficacy model M4 with covariates on $\varepsilon$ | Age (> 65 years) | -0.84 |
| | | Vaccination (≥ 2 doses) | 1.55** |

<sup>a</sup> Candidate models used for sensitivity analyses, can refer to **Supplementary Table S2**.
<sup>b</sup> Model Estimate ( $\beta$ ) is the modeled covariate effect on target parameters. For covariates on EC<sub>50</sub> (**M5, M6**), positive values indicate an increase in EC<sub>50</sub> (reduced treatment efficacy), and negative values indicate a decrease in EC<sub>50</sub> (enhanced treatment efficacy). Conversely, for covariates on constant efficacy $\varepsilon$ (**M7, M8**), positive values indicate an increase in overall efficacy (enhanced treatment), and negative values indicate a decrease in efficacy (reduced treatment).
<sup>c</sup> \*p < 0.05; \*\*p < 0.01; \*\*\*p < 0.001

