## Supplementary figures and images for "Host Factors Modulate Nirmatrelvir–Ritonavir Efficacy in COVID-19 Patients: A Viral Dynamics Modeling Study"

### Supplementary Figure 1

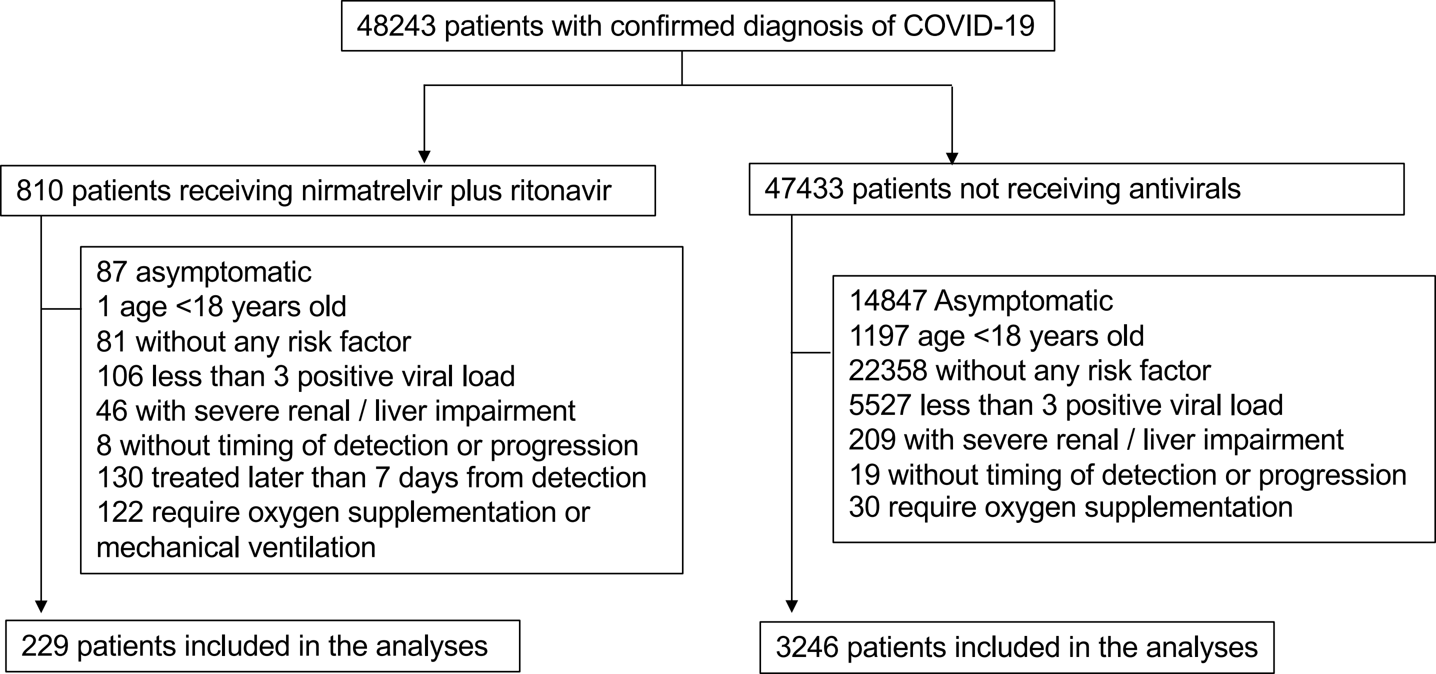

### Supplementary Figure 2

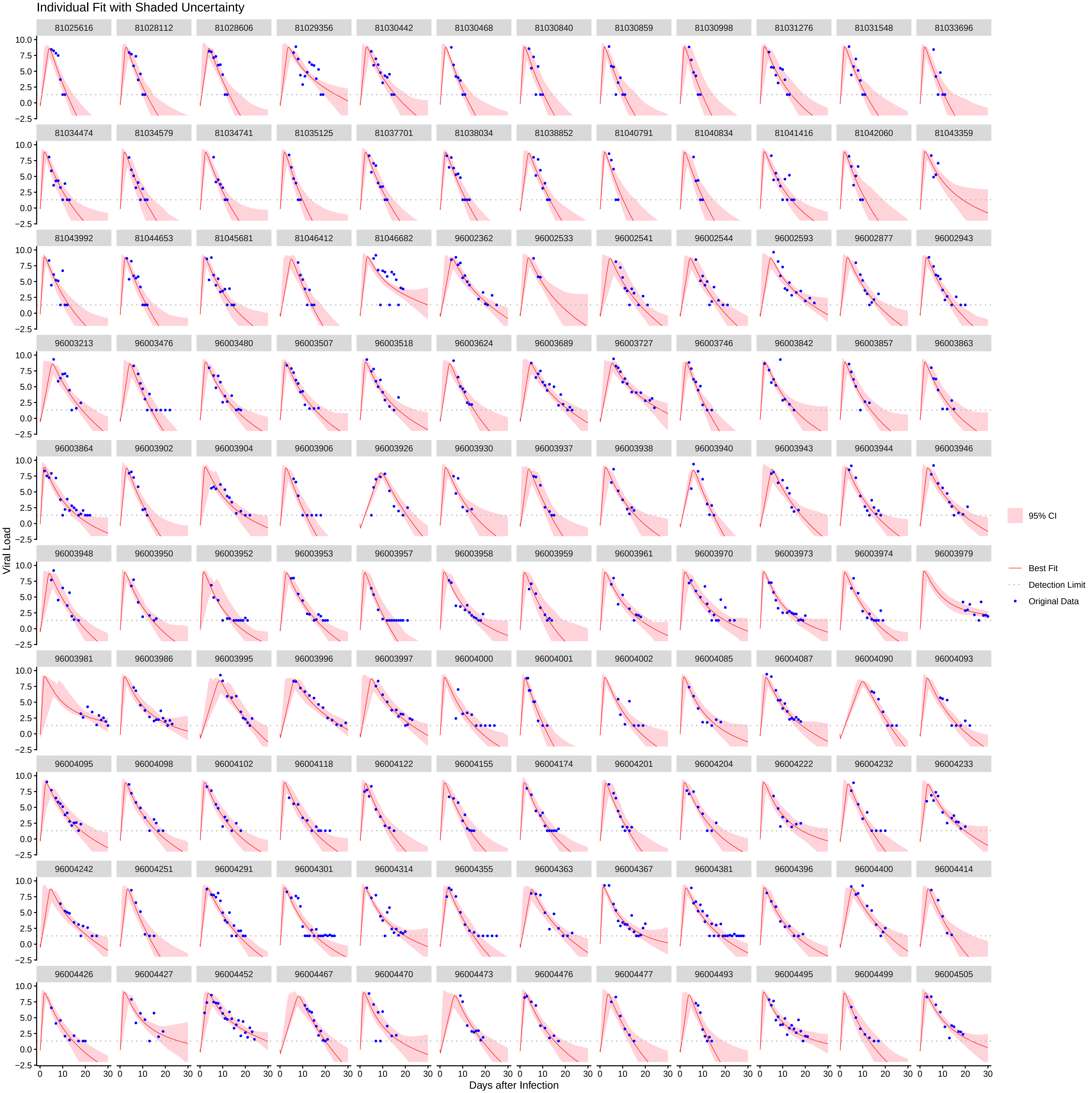

Individual Fit with Shaded Uncertainty

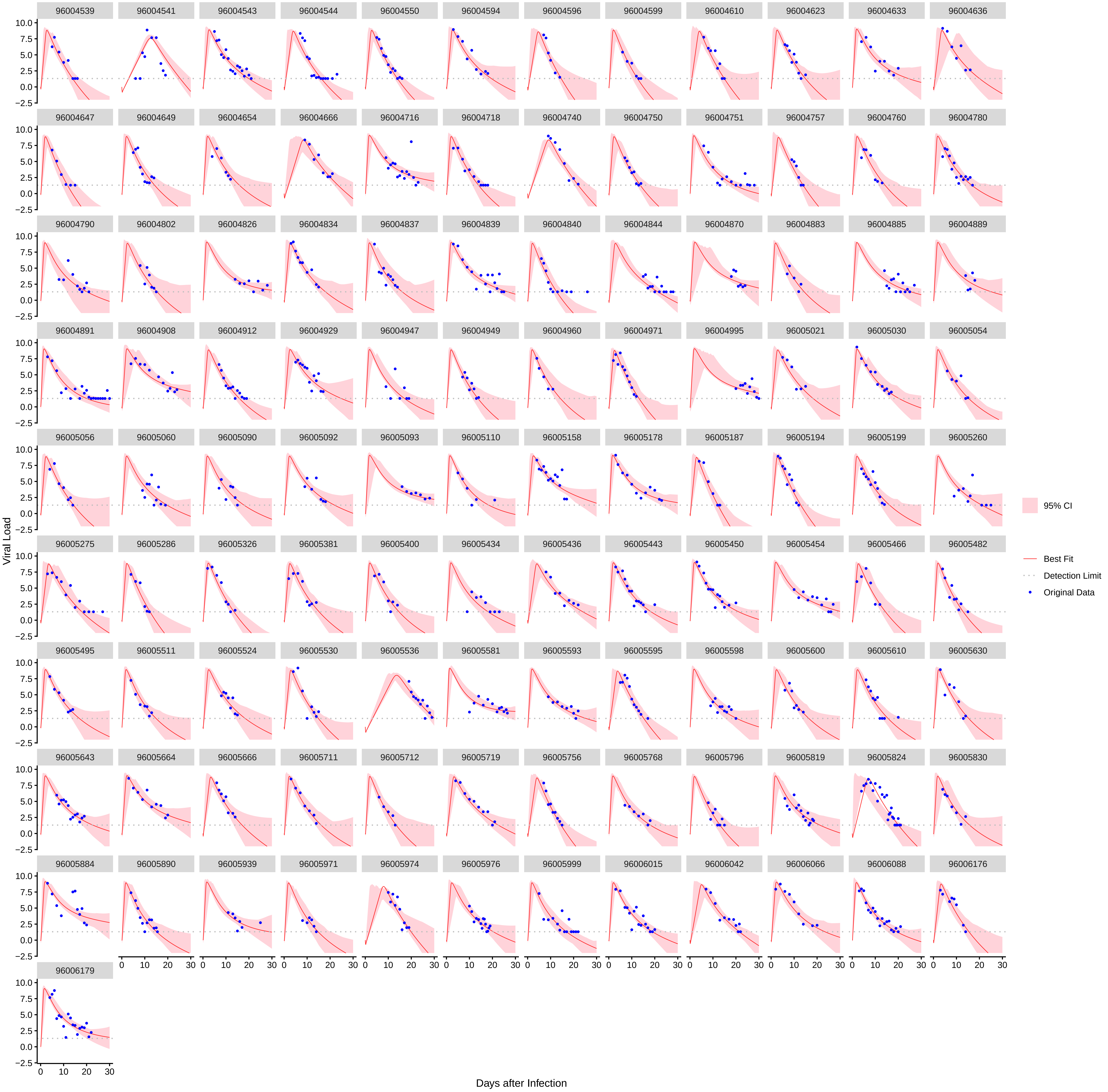

### Supplementary Figure 3

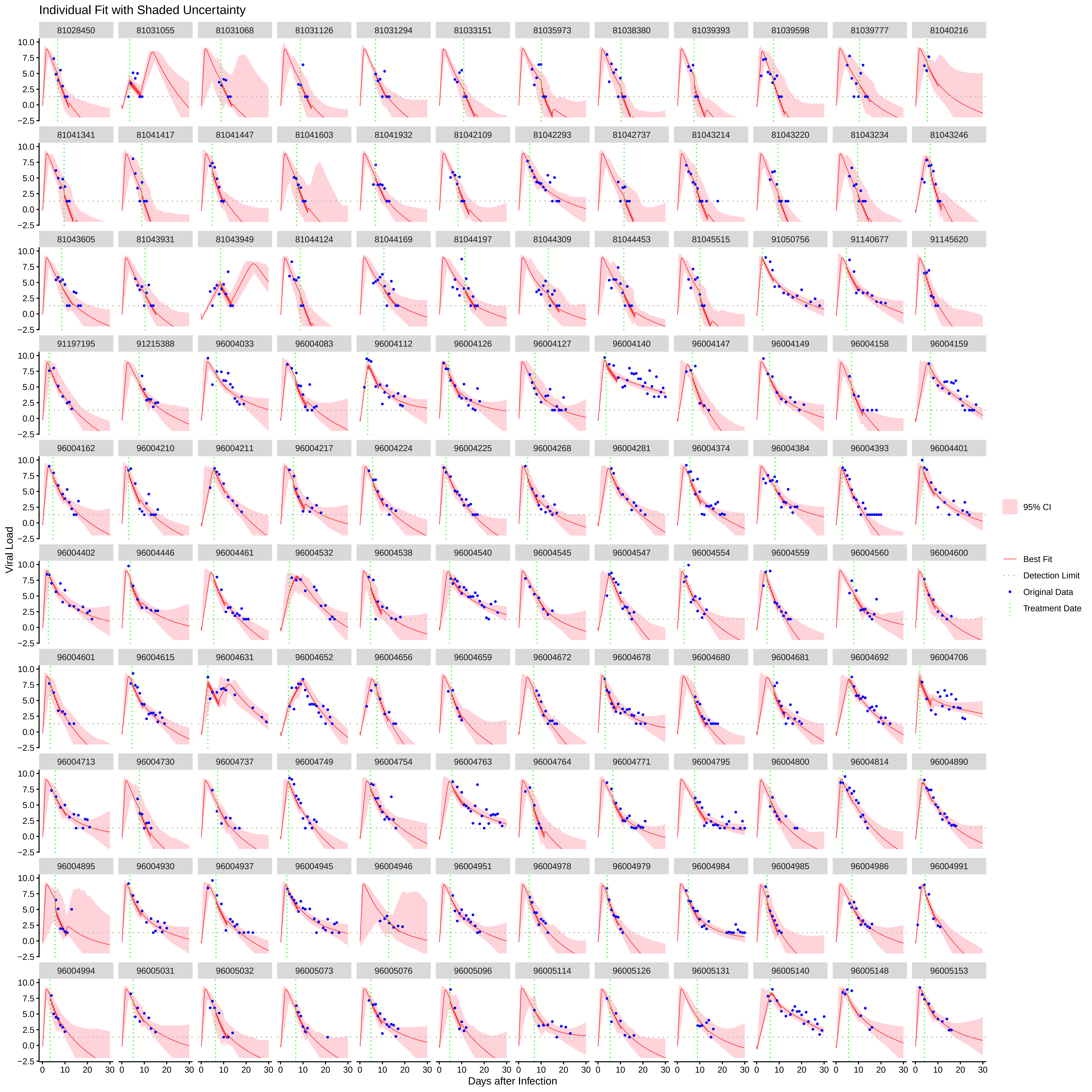

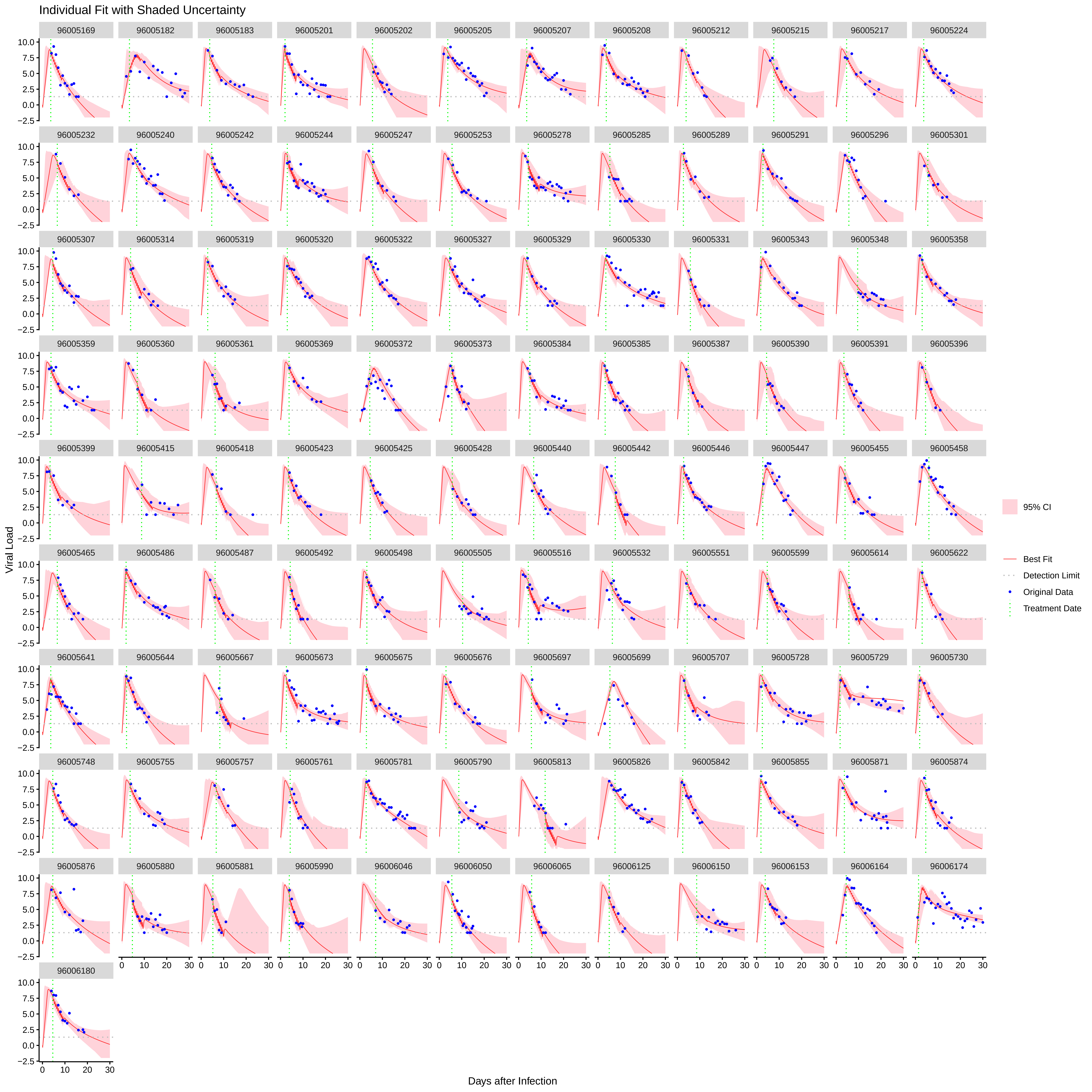

### Supplementary Figure 4

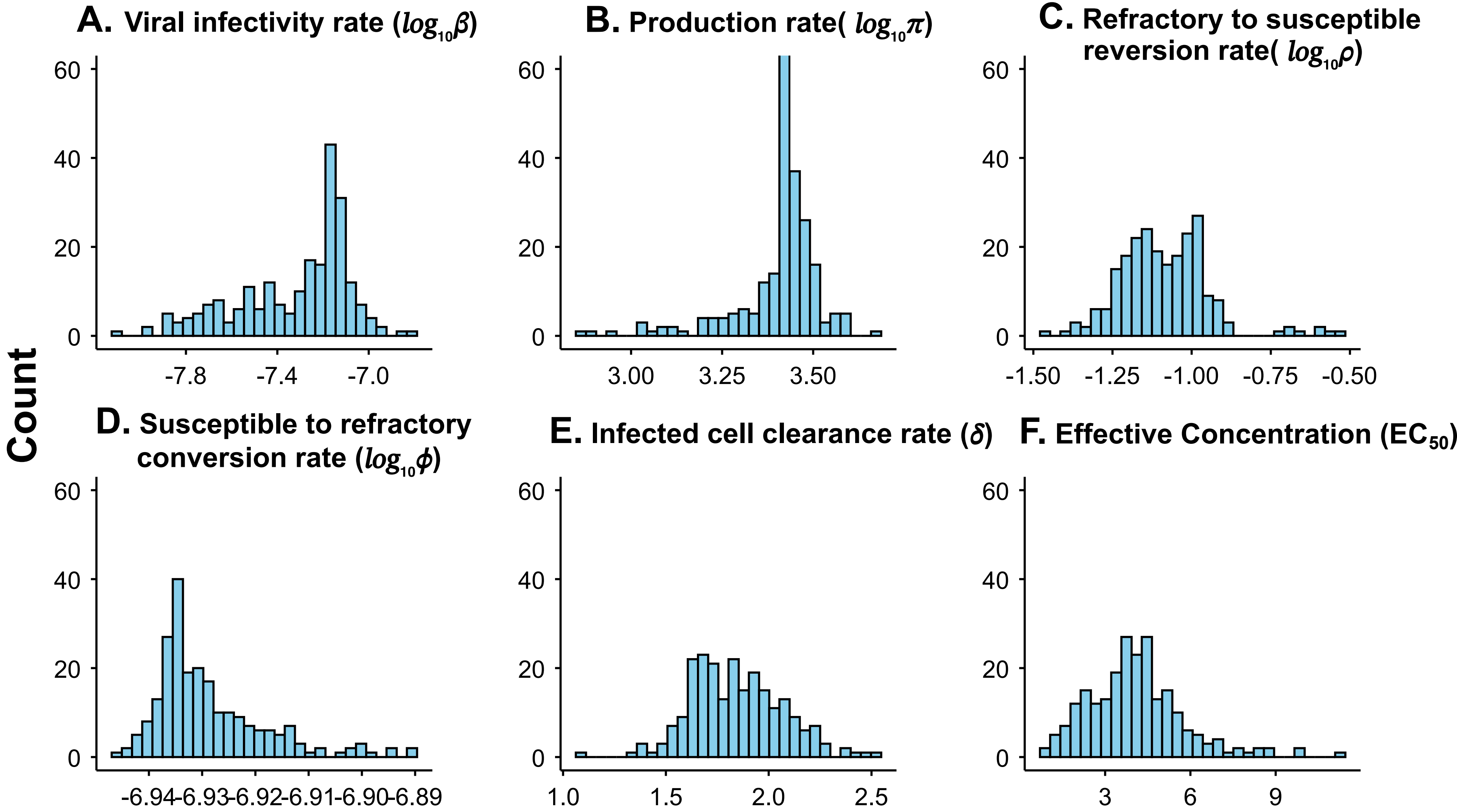

### Supplementary Figure 5

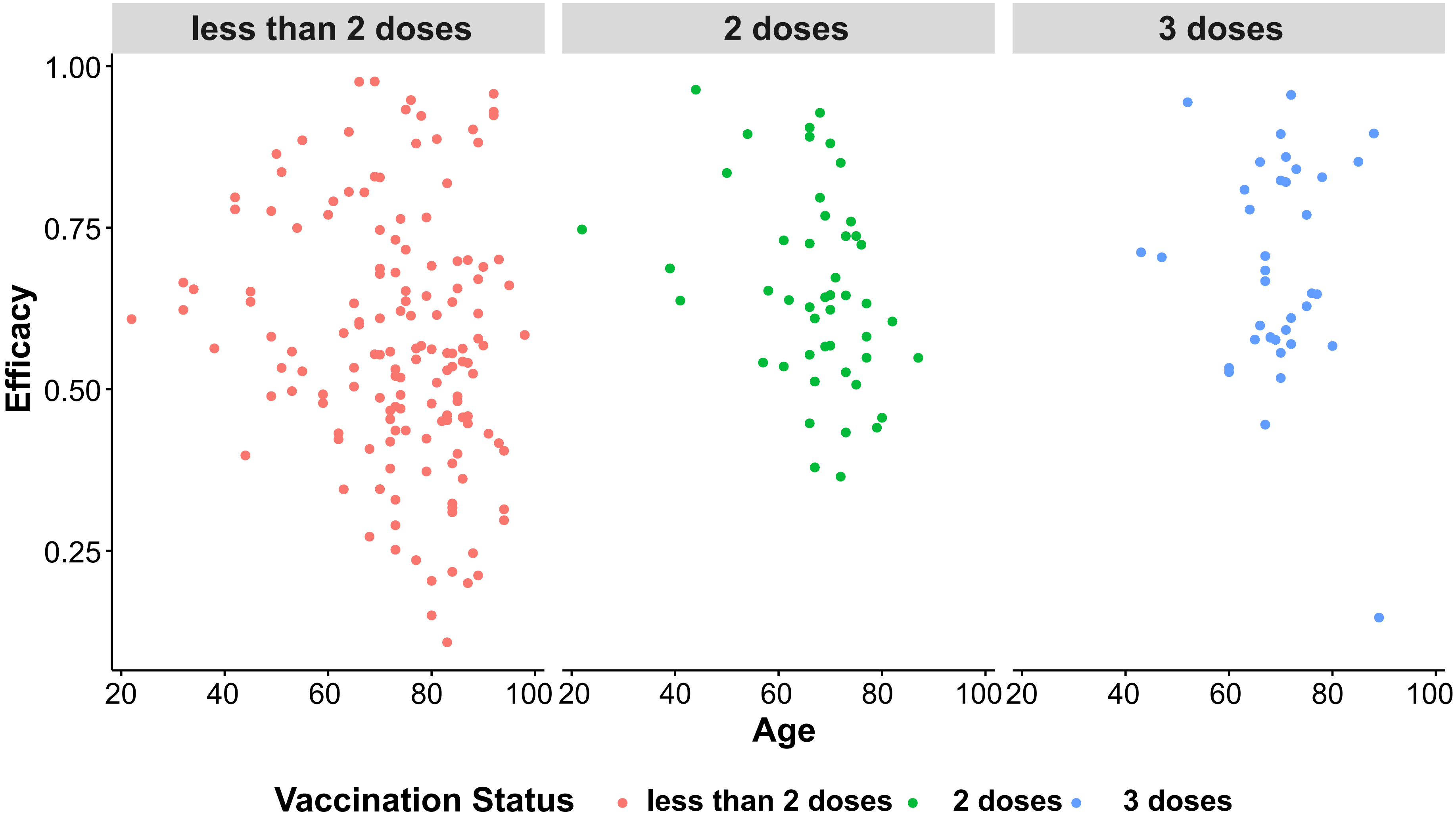

### Supplementary Figure 6

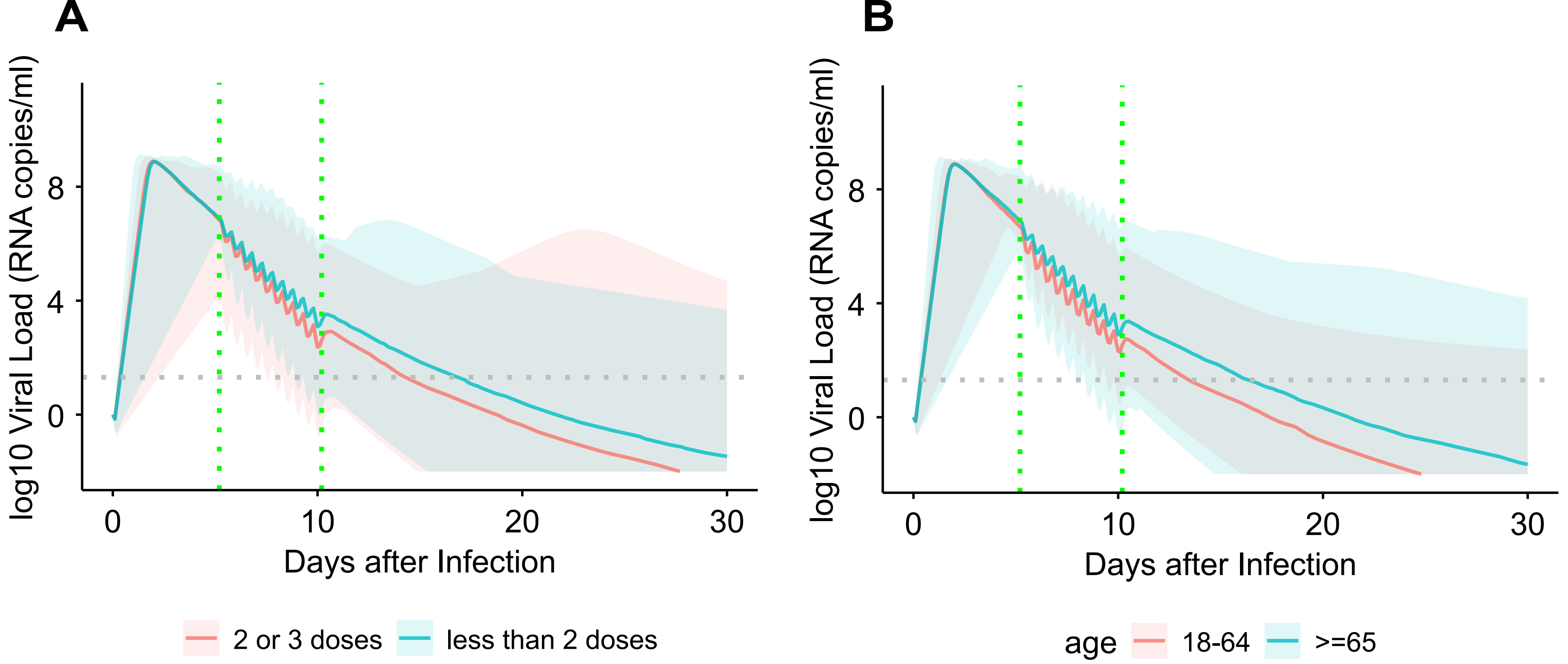
